# Validation of case correctness and time intervals agreement in the Swedish registry of cardiopulmonary resuscitation using emergency medical dispatch data, 2015–2024

**DOI:** 10.64898/2026.02.20.26346753

**Authors:** Erik Boberg, Carl Magnusson Martin, Douglas Spangler, Fredrik Byrsell, Martin Jonsson

## Abstract

**Objective:** To validate case number correctness and time interval agreement in the Swedish Registry of Cardiopulmonary Resuscitation (SRCR) for out-of-hospital cardiac arrest (OHCA) by linkage to Emergency Medical Dispatch Centre (EMDC) data between 2015 and 2024.

**Methods:** In this retrospective validation study, OHCA records reported to the SCRC were linked with EMDC-indexed OHCA for validation and correction of EMS case numbers. We quantified the proportion of correct EMS case numbers reported as agreement for fully correct and partially correct EMS case numbers in SRCR. Time interval agreement was assessed by comparing dispatch to arrival (unit response time) and call start to arrival (total response time) between SRCR and EMDC. For each linked case, time differences were calculated as (SRCR – EMDC) in seconds. Median differences were estimated using Bayesian quantile regression.

**Results:** EMS case number completeness was high, but the proportion of fully correct case numbers was limited. Among 56,969 SRCR records, 1,004 (1.8%) lacked an EMS case number. The proportion of SRCR records with partially correct EMS case numbers was around 90% up to the year 2020 and declined to 85% in 2022–2024. Dispatch-related time intervals showed high agreement between sources, with a median difference of −0.3 seconds (95% CrI –3.9 to 4.0). In contrast, SRCR total response time (from dispatch call answer to arrival at scene) was shorter than EMDC, with a median difference of 80.9 seconds (95% CrI –84.7 to –77.0).

**Conclusion:** SRCR unit response time reflects EMDC operational recording. The SRCR total response times were consistently shorter than the interval at the EMDC, indicating a potential underestimation of the total EMS response time in the registry.

## Introduction

Out-of-hospital cardiac arrest (OHCA) remains a leading cause of death in Europe.^1^ Survival is strongly time-dependent,^2^ relying on early recognition, bystander cardiopulmonary resuscitation (CPR), defibrillation, and rapid emergency medical services (EMS) response.^3,4^

The American Heart Association (AHA) and European guidelines promote cardiac arrest registries as a cornerstone of quality improvement.^5^ Sweden is one of nine European countries that have an OHCA registry covering the entire population.^6^ Notably, an often-cited 80% coverage estimate for the Swedish Registry of Cardiopulmonary Resuscitation (SRCR)^7^ is derived from local audits rather than from a national validation against an external reference source. A regional Swedish validation study reported that approximately one-quarter of OHCA cases were not reported prospectively by EMS, suggesting potential selection bias in registry-based analyses.^8^ The SRCR, established in 1990, functions as a national quality registry for all OHCA.^9^ Because healthcare is delivered across 21 regions in Sweden with multiple EMS providers, a national registry is essential to enable surveillance and benchmarking of outcomes across the chain of survival.^10^

The usage of SRCR-based estimates depends on case ascertainment and accurate recording of Utstein variables, including EMS time stamps. Time intervals are frequently used to evaluate system performance and as covariates in outcome modelling. In the most recent Utstein guidelines, the importance of EMS time variables is highlighted, and the recommended gold standard response interval is from when the dispatch centre answers the call to EMS arrival at the scene.^11^

Registration accuracy is not static and may change over time as EMS systems evolve and documentation and workflows are updated. Without contemporary validation against an external reference, systematic errors may go undetected, affecting regional comparisons, temporal trend analyses, and research findings. Emergency medical dispatch centre (EMDC) data provide operational time stamps recorded in real time and are therefore well-suited as a reference for validating EMS time variables reported in the SRCR. To our knowledge, no comprehensive validation of SRCR case ascertainment and time agreement against EMDC data has been performed.

This study aims to investigate the quality of reported case numbers and agreement between time intervals (unit response time and total response time) in the SRCR and the EMDC.

## Methods

### Study design and setting

This retrospective validation study compared SRCR OHCA records with EMDC-indexed OHCA records as an external reference. The study period was January 1, 2015, to December 31, 2024. Ethical approval was granted by the Swedish Ethical Review Authority (Dnr 2025-03753-01). Analytic data and materials, together with the code required to reproduce the analysis and figures, are available at Open Science Framework (URL)

### Data Sources

#### The Emergency Medical Service organization

OHCA reporting to the SRCR follows Utstein-based definitions and includes key operational timestamps, such as dispatch receipt and EMS arrival. These timestamps may be system-generated or entered manually by EMS personnel, depending on regional workflows and infrastructure. Thereafter, OHCA data are submitted to the SRCR either through direct entry into a secure electronic cardiac arrest report template or through automated transfer from the regional EMS patient record system.

#### The Emergency Medical Dispatch Centers

Sweden uses a centralized public safety answeing point (PSAP) managed by SOS Alarm, which manages all emergency calls nationwide. SOS Alarm also manages EMDC operations for 17 out of Swedens 21 regions, while four instead use an independent EMDC organization, Sjukvårdens Larmcentral (SvLc). In SvLc regions, EMDC is integrated with the EMS organization. If a patient is reported as unconscious with abnormal breathing, all Swedish EMDC indexes the incident as a suspected cardiac arrest. Dispatch data and audio recordings are stored separately from EMS clinical records, and there is no automated linkage between EMDC and the SRCR.

#### The Swedish Registry of Cardiopulmonary Resuscitation

Participation in SRCR is voluntary, and all Swedish EMS organisations report OHCA to the registry. The registry is hosted by the Centre of Registries, Västra Götaland, which provides technical and legal support for registry infrastructure and governance. The registry includes all OHCA where cardiopulmonary resuscitation (CPR) was initiated, and since 2010, all EMS agencies have taken part in the registry. SRCR data are collected using a standardized resuscitation template^12^ based on the AHA Utstein recommendations^11^. Through annual reports, the SRCR describes temporal trends in resuscitation care and provides comparisons across healthcare regions. Survival at 30-days is obtained automatically through linkage with the Swedish Population Registry.

#### Study population

All SRCR records registryed as OHCA, together with all EMDC-indexed suspected OHCA between January 1, 2015, and December 31, 2024, were extracted. No exclusion criteria were applied.

#### Study Objective

The primary objective was to quantify the proportion of SRCR OHCA records with a correctly recorded EMS case number, using EMDC as the reference and distinguishing between case numbers that were fully correct and partially correct. We also assessed agreement in EMS dispatch and arrival time intervals by comparing EMDC-recorded time intervals with the corresponding SRCR intervals.

#### Record linkage

We linked SRCR OHCA records to EMDC suspected-OHCA events by hierarchical matching. First, we linked all exact case numbers. Second, we used partial case-number matching to account for common formatting variations (missing prefixes or suffixes). Third, for SRCR records with missing, incomplete, or non-linkable case numbers, we performed a manual review using event date, timestamps, municipality, and region to identify the most plausible EMDC record. A case was considered a match when the SRCR OHCA and an EMDC suspected-OHCA call occurred on the same date and region within a narrow time window (e.g., SRCR time 02:01 and an EMDC call logged at 02:00 in Stockholm County).

Case-number formats also varied by dispatch organisation. For SOS Alarm, the case identifier comprises a one or two-digit dispatch centre code, followed by a 6–8 digit sequential number. Together, they form a unique case. Some records also include a suffix if additional resources were dispatched (e.g., −2). For Sjukvårdens Larmcentral (SvLc), the identifier consists of the date in YYYYMMDD format, followed by a three-digit region code, and a 1–3 digit component representing the serial number of the assignement over the 24 hour period. Examples of correct EMDC formats and common SRCR variations are provided in Table 1.

**Table 1.** Examples of corrected EMDC case number formats and common SRCR entry variants. The case-number structure used in EMDC systems (SOS Alarm and SVLC) and common variants observed in SRCR illustrate how EMS personnel recorded case numbers in the registry. The table also shows if the SRCR variant supports a partial or complete match for case ascertainment. EMDC = Emergency Medical Dispatch Center, SRCR = Swedish Registry of Cardiopulmonary Resuscitation.

| EMDC | Correct case number | Recorded in SRCR | Partial match<sup>*</sup> | Fully match<sup>†</sup> |
| --- | --- | --- | --- | --- |
| **SOS alarm** | 19-9876543 | 19-9876543 | √ | √ |
|  |  | 19-98765432-002 | √ | √ |
|  |  | 1998765432 | √ | √ |
|  |  | 98765432-2 | √ | X |
|  |  | 98765432 | √ | X |
|  |  | 97654322 | X | X |
| **SvLc** | 20260101-100-26 | 20260101-100-26 | √ | √ |
|  |  | 100-20260101-26-1 | √ | √ |
|  |  | 20260101-100-26-1 | √ | √ |
|  |  | 10020260101261 | √ | √ |
|  |  | 20260101-26 | X | X |
|  |  | 20260101-100 | X | X |

\* Sufficient information to reconstruct and match a unique EMDC case number.
† Sufficient information to identify a match.

#### Calculation of time intervals in SRCR and EMDC

After correction and linkage of EMS case numbers, we calculated the response time interval from each data source. In SRCR, the total response time interval was calculated as the time from call registration (AR_AlarmRegT) to EMS arrival at the scene (AR_AmbArrStopT). The unit response interval was calculated as the time from EMS dispatch (AR_AlarmOutT) to EMS arrival at the scene (AR_AmbArrStopT). In the EMDC data, the total response interval was calculated as the time from call start (Created) to EMS arrival at the scene (Status_F), and the unit response interval was calculated as the time from EMS dispatch (Status_T) to EMS arrival at the scene (Status_F). All intervals were calculated in seconds and the difference between the two sources was compared as (SRCR – EMDC)

### Statistical analysis

The proportion of correctly registryed OHCA cases in the SRCR is reported as the percentage of agreement between the originally recorded EMS case number and the corrected case number after manual correction. Agreement is reported both for fully and partially corrected case numbers

To quantify differences in dispatch and arrival time reporting between SRCR and EMDC, we modelled the differences in time intervals using Bayesian quantile regression with an asymmetric Laplace likelihood. Analyses were performed in brms.^13^ For each time interval, we fitted intercept-only models to estimate the population-level median for SRCR and EMDC separately. We used a normal prior for the intercept with a mean of 600 seconds and a standard deviation of 200 seconds. The prior can be translated to a median response time of somewhere between 3.3 and 16.7 minutes. The median difference was then derived by subtracting posterior draws from the EMDC model from those of the SRCR model (SRCR – EMDC), yielding a posterior distribution for the median difference. All analyses were repeated by calendar year and by healthcare region. Results are reported as the posterior median difference with 95% credible intervals (CrI), together with the full posterior distribution.

## Results

Between 2015 and 2024, 57,708 OHCA were reported to the SRCR. After removal of duplicate entries, 56,969 remained for analysis. Among these, 1,004 (1.8%) had no recorded EMS case number.

### Valid EMS case number by healthcare region

The proportion of fully correct EMS case numbers varied by healthcare region. Two regions had 0% fully corrected case numbers, whereas Stockholm had the highest proportion of fully corrected case numbers at 98%. For partially correct case numbers (missing dispatch center code), the proportion also varied by region, ranging from 77% in Central Sweden to 100% in Stockholm (Figure 1).

**Figure 1.**
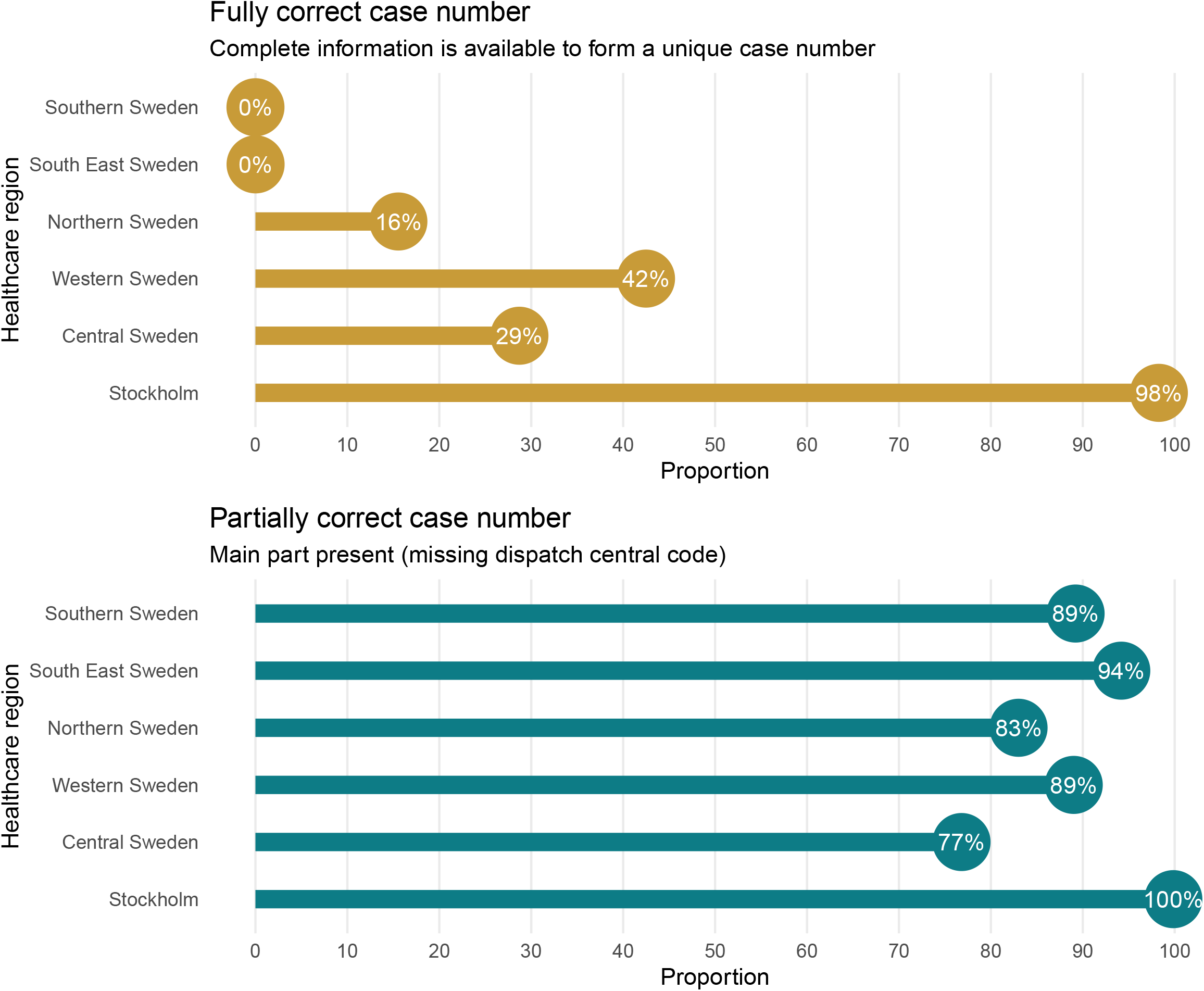
Proportion of SRCR records with fully and partially correct EMS case numbers, by healthcare region. Proportion of SRCR records with (A) fully correct and (B) partially correct EMS case numbers, by healthcare region. A fully correct EMS case number indicates a complete case number suitable for direct linkage to EMDC records. A partially correct case number indicates that the main case-number component is present, but the dispatch central code is missing, requiring correction before linkage. SRCR = Swedish Registry of Cardiopulmonary Resuscitation, EMDC = Emergency Medical Dispatch Center, EMS = Emergency Medical Services.

### Valid EMS case number per year

The proportion of SRCR records with partially correct case numbers was approximately 90% until 2020, but declined in later years to around 85% in 2022–2024. The proportion of records with a fully correct EMDC case number increased from 32% to 37% during 2015-2019, to above 40% during 2020-2024 (Figure 2).

**Figure 2.**
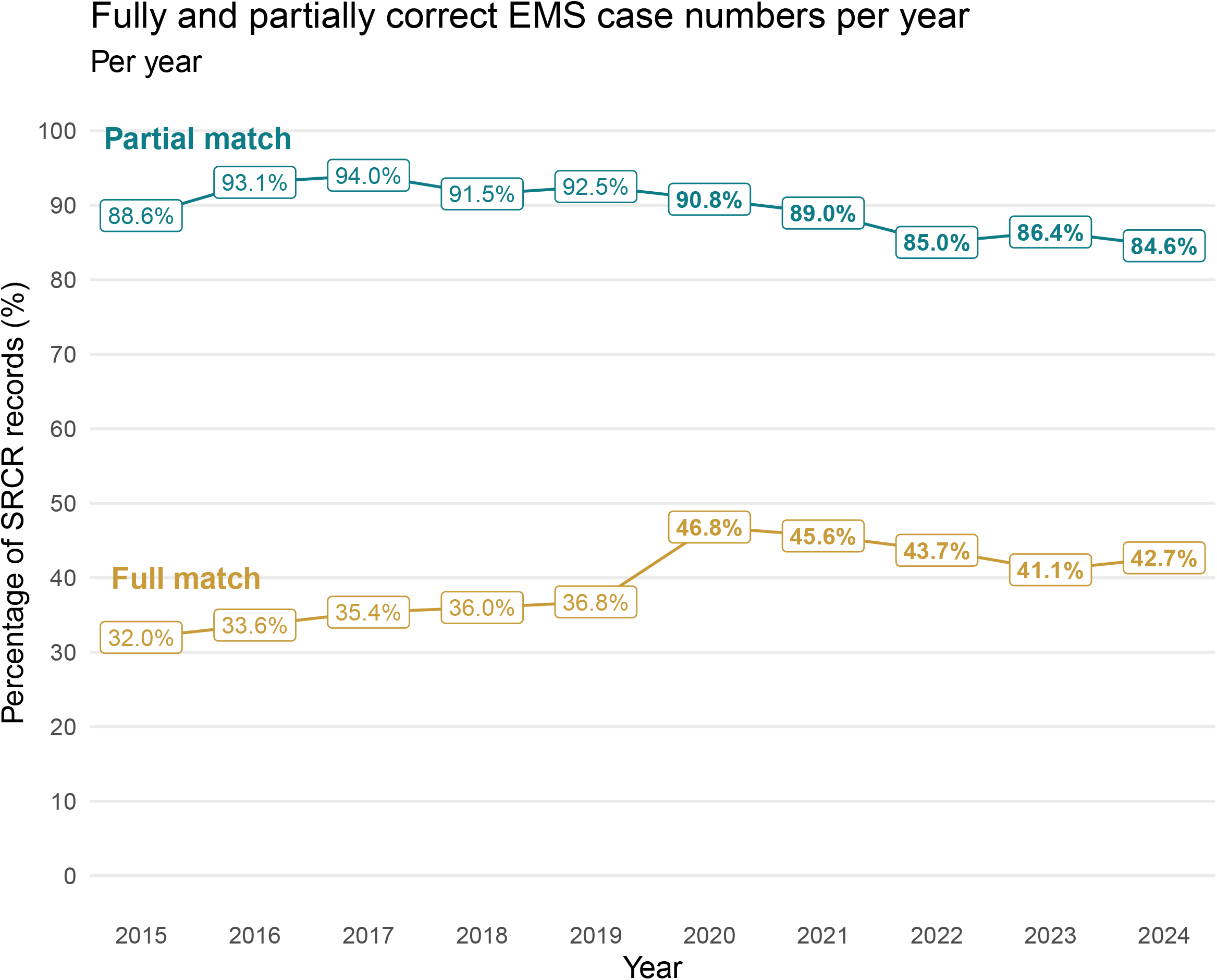
Temporal trends in fully and partially correct EMS case numbers recorded in the SRCR, 2015–2024. Proportion of SRCR OHCA records with fully correct and partially correct EMS case numbers by year (2015–2024). The figure shows consistently high levels of partially correct case numbers through 2020, followed by a modest decline in later years, alongside an increase in fully correct case numbers after 2020. SRCR = Swedish Registry of Cardiopulmonary Resuscitation, OHCA = Out-of-hospital cardiac arrest, EMS = Emergency Medical Services.

### Difference in response interval

For the unit response interval (EMS dispatch receipt to arrival at scene), the median difference between SRCR and EMDC was −0.3 seconds (95% CrI = −3.9 to 4.0). For the total response interval (dispatch call answer to arrival at scene) SRCR time was shorter than EMDC, with a median difference of 80.9 seconds (95% CrI = −84.7 to −77) (Figure 3).

**Figure 3.**
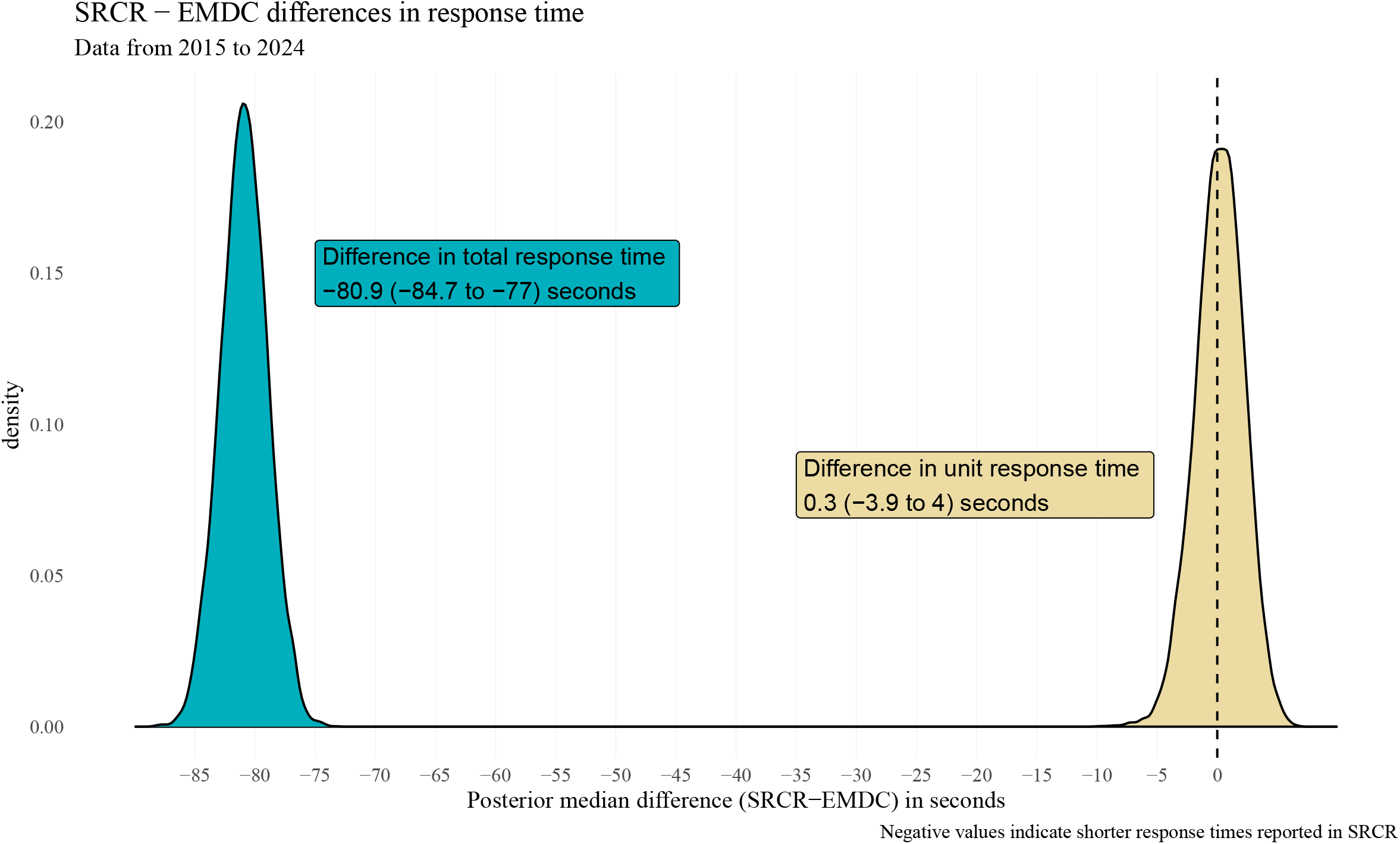
Systematic differences in unit and total response time between SRCR and EMDC (2015–2024). Posterior distributions of the median difference in response times between SRCR and EMDC for 2015–2024. The x-axis represents the difference in seconds (SRCR – EMDC). Negative values indicate that the response times reported in SRCR are shorter than those in EMDC. The total response interval (dispatch call answer to arrival at scene) shows a clear negative shift with a median difference of –80.9 s. In contrast, the unit response interval (EMS dispatch receipt to arrival at scene) is centered near zero, indicating a negligible systematic offset. SRCR = Swedish Registry of Cardiopulmonary Resuscitation, EMDC = Emergency Medical Dispatch Center, EMS = Emergency Medical Services.

Regional analysis showed that the unit response interval differences varied across healthcare regions. The smallest difference was observed in Stockholm (−4 seconds, 95% CrI = −14 to 8), followed by Central Sweden (−20 seconds, 95% CrI =-29 to −11.4) and Western Sweden (−51 seconds, 95% CrI = −58 to −43). The biggest discrepancy was observed for Southern Sweden (−74 second, 95% CrI = −82 to −60).

For the total response interval, discrepancies were larger. Stockholm showed the least evidence of systemic difference (median – 32 seconds, 95% CrI = −40 to 26), whereas all other regions had median differences exceeding 70 seconds, with the largest difference in Central Sweden (−132 seconds, 95% CrI=-142 to −141) (Figure 4).

**Figure 4.**
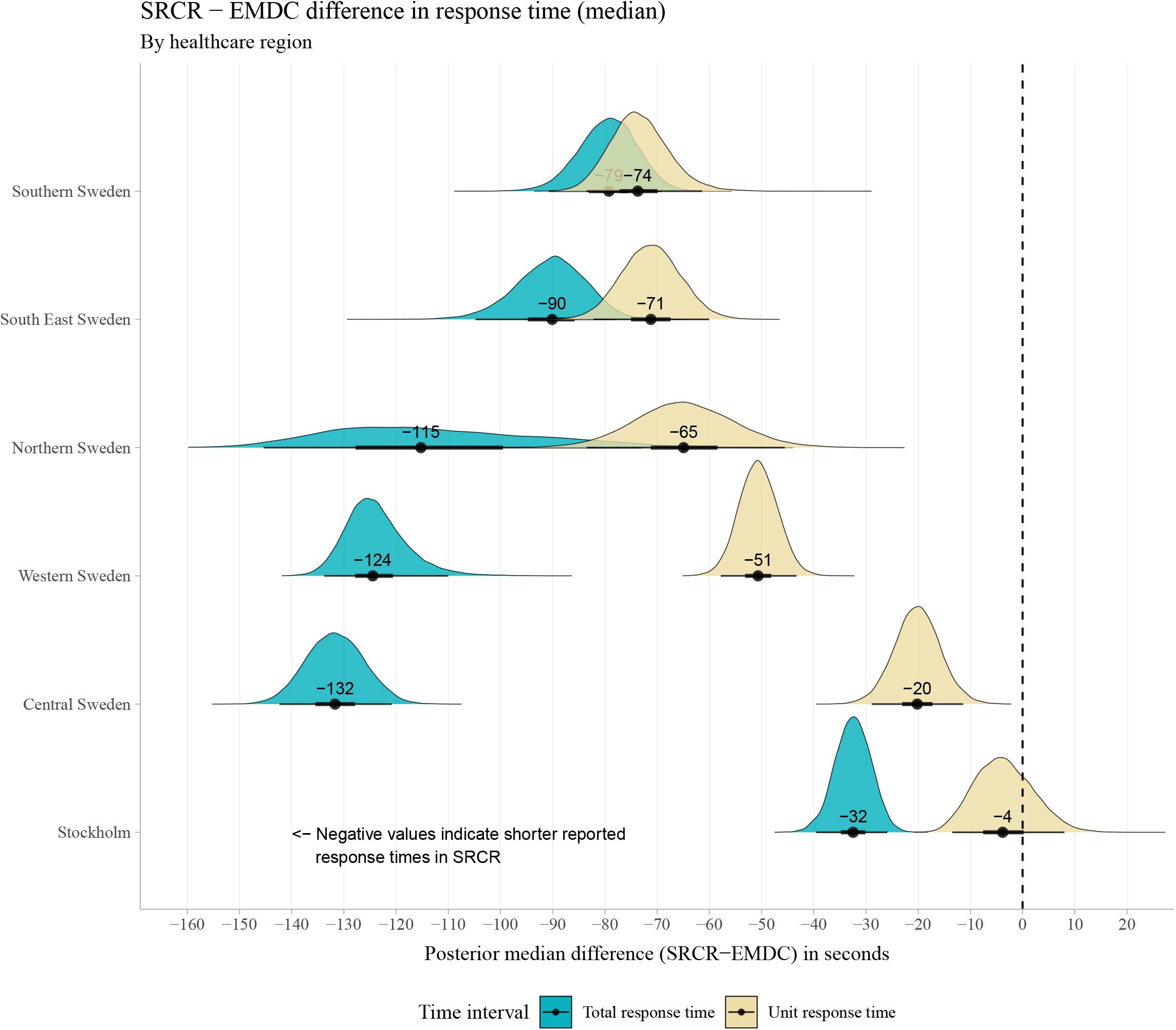
EMDC–SRCR differences in total and unit response time by healthcare region. Posterior distributions of the median difference in response times between SRCR and EMDC, by healthcare region. The x-axis shows SRCR–EMDC in seconds; negative values indicate shorter response times reported in SRCR compared to EMDC. Teal distributions represent the total response time (dispatch call answer to arrival at scene), and Beige distributions represent the unit response time (EMS dispatch receipt to arrival at scene). Points and horizontal lines indicate the posterior median and 95% credible interval for each region. SRCR total response times are systematically shorter than EMDC across all regions, whereas unit response times show smaller and more variable differences. SRCR = Swedish Registry of Cardiopulmonary Resuscitation, EMDC = Emergency Medical Dispatch Center, EMS = Emergency Medical Services.

Year-specific estimates are shown in Figure 5. The median unit response interval difference ranged from −42 seconds in 2019 (95% CrI = −62 to −1) to close to zero in 2020. For the total response interval, the largest discrepancy occurred in 2015 when SRCR response times were more than 2 minutes shorter than EMDC (95% CrI = −146 to −119). The smallest difference occurred in 2024 (−67 seconds, 95% CrI = −80 to −54).

**Figure 5.**
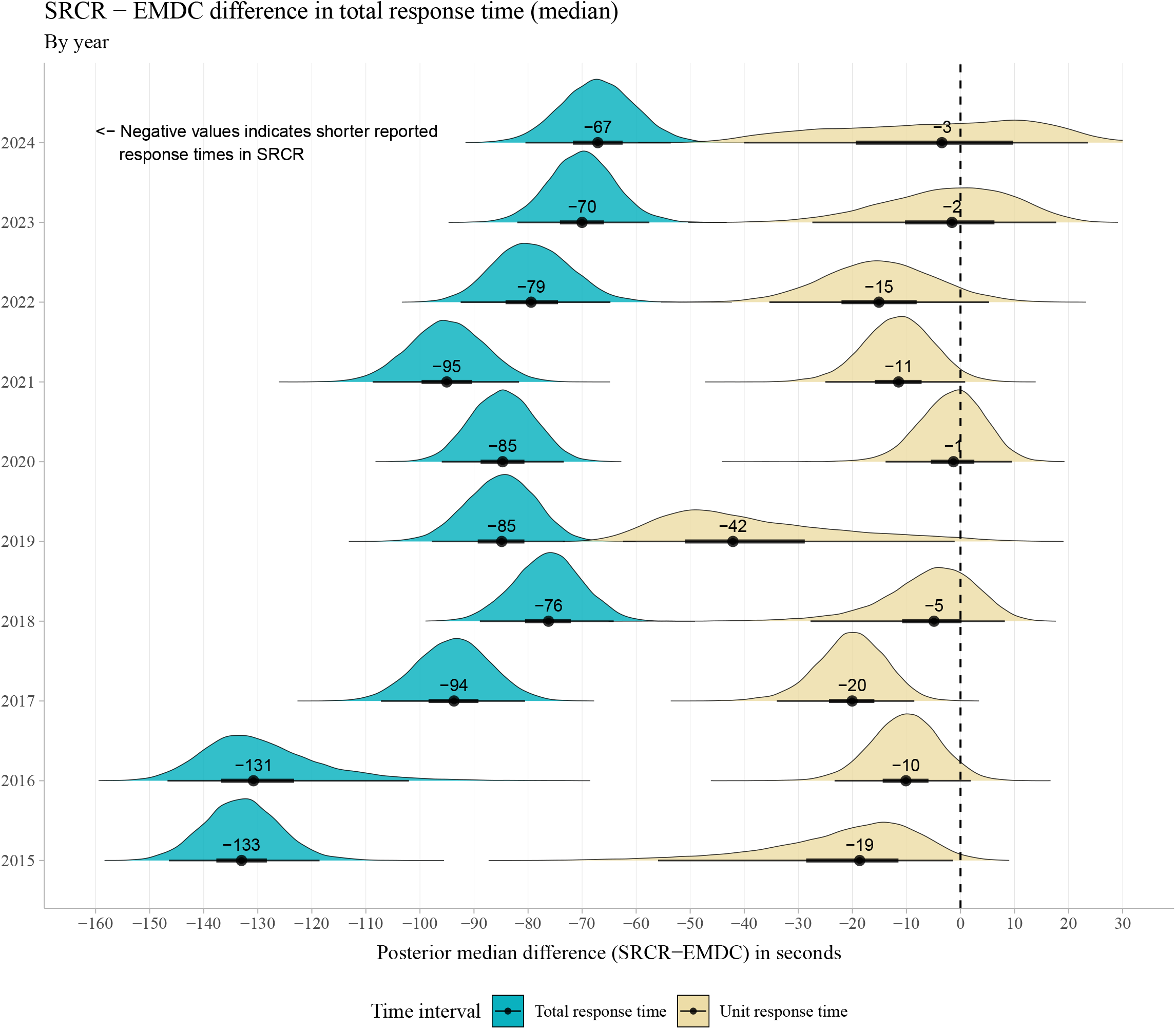
Yearly differences in total and unit response time between SRCR and EMDC (2015–2024). Posterior distributions of the yearly median difference in response times between SRCR and EMDC, 2015–2024. The x-axis shows SRCR–EMDC in seconds; negative values indicate shorter response times reported in SRCR than in EMDC. Teal represents the total response time (dispatch call answer to arrival at scene), and beige represents the unit response time (EMS dispatch receipt to arrival at scene). For each year, the point and horizontal line show the posterior median and 95% credible interval. Across all years, total response time is systematically shorter in SRCR than EMDC, whereas unit response time shows smaller and often near-zero differences. SRCR = Swedish Registry of Cardiopulmonary Resuscitation, EMDC = Emergency Medical Dispatch Center, EMS = Emergency Medical Services, SRCR.

## Discussion

This validation study reports SRCR case correctness and agreement in EMS time intervals using EMDC-indexed OHCA events as an external reference. By linking SRCR records to EMDC events through dispatch case numbers, by structured matching of time, date, municipality, and region, we were able to quantify the proportion of SRCR cases that could be reliably identified and to evaluate concordance between SRCR and EMDC time reporting for EMS dispatch and arrival at the scene.

A central finding was that successful linkage depended on the quality of EMS case numbers recorded in SRCR. Because the EMDC case number is system-generated, linkage failures predominantly reflected SRCR data quality, including missing case numbers, inconsistent formatting, or transcription errors. This is not only relevant for routine registry validity but also has implications for OHCA research and quality improvement, since incorrect case numbers impede linkage to other external data sources that rely on the same identifier, such as the Swedish community volunteer responders system or EMS charts, with the potential to reduce sample size or introduce selection bias.

The high proportion of fully correct case numbers in Stockholm likely reflects a more automated workflow. In this healthcare region, SRCR reporting is integrated into routine EMS documentation. An OHCA patient record cannot be finalized without submission of the SRCR report to their local EMS system. This type of forced completion likely improves identifier quality compared with manual or retrospective reporting

The Utstein guidelines^11^ recommend measuring the total response time using the time from call answering at the PSAP to EMS arrival at the scene (total response time). We found a large discrepancy between the total response time reported in SRCR and that recorded in the EMDC, suggesting a consistent offset. To address this, a routine linkage between Utstein registrys and the EMDC should be considered to enhance the quality of cardiac arrest registries. Linkage between registries and the EMDC could also ease the burden on EMS personnel, as they do not have to fill in time stamps.

It is worth commenting on the finding that the unit response time seem to be very similar between the SRCR and the EMDC when all data are pooled, but when reported by year/region differences appear. This is due to the skewed distribution of the time variables. What happens is very similar to what is known as Simpson’s paradox.

As the format of timestamps reported into the SRCR is HH:MM, there is a high risk of rounding. Rounding could shorten recorded intervals and could make EMS response times appear faster, which is important when registry data are used for annual reports or observational analyses. We believe this pattern is observed because EMDC times are generated automatically in real time within the dispatch platform. In contrast, SRCR timestamps may be derived from EMS documentation that often involves manual entry, retrospective patient reports, or local system differences. Additionally, OHCA incidents may be attended by multiple EMS units, and the unit completing the SRCR report may not be the first unit on scene. In these cases, the recorded time of arrival at the scene in SRCR may correspond to the reporting unit’s arrival instead of the earliest EMS arrival, which could contribute to discrepancies when compared with EMDC times.

Future registry improvements could include automated arrival and departure times using GPS-based geofencing. If OHCA events were assigned a geographic coordinate, EMS vehicle positions could be monitored in real time. Arrival at the scene, defined as the first timestamp at which the unit enters a predefined geofence around the OHCA location, and leaving from the scene, defined as the timestamp at which the EMS unit exits the geofence. Such automated time stamps could reduce rounding and manual-entry errors.

These findings have direct implications for both registry governance and research use. For researchers, SRCR is suitable for epidemiologic and quality improvement analyses, but EMS time variables should be treated with care, particularly for response time analyses. For SRCR, standardising the EMS case-number format and integrating EMDC data into SRCR would improve linkage, increase sample size, and support routine validations.

## Strengths

A major strength of this study is the use of a large national EMDC dataset for linkage and validation, combined with a transparent, hierarchical case number matching. Another strength is focusing on EMS time stamps that are clinically important and frequently used in OHCA research.

## Limitations

This work should be interpreted in light of several limitations. First EMDC data do not provide a population denominator for incidence, and we could not estimate national SRCR case completeness because the number of EMS-treated OHCAs missing from SRCR is unknown. Second, although EMDC time stamps are system-generated during call handling, they can still contain recording errors (e.g., premature registration of arrival at the scene). Therefore, agreement should be viewed as concordance between two routinely collected data sources. Finally, manual linkage may introduce a small risk of incorrect matches, especially in rare circumstances with multiple OHCA events in close temporal proximity within the same region.

## Conclusion

SRCR unit response time showed high agreement with the EMDC. In contrast, SRCR total response time was systematically shorter than EMDC. This has practical value for the interpretation since reported times may be underestimated. Standardising the EMS case-number format and integrating EMDC data into SRCR could increase the validity of national registry data.

## Data Availability

Analytic data and materials, together with the code required to reproduce the analysis and figures, are available at Open Science Framework (URL). Data will be made public if accepted in circulation, below is view-only link for reviewers and editor.

https://osf.io/dqja5/overview?view_only=7ad2d890f4aa406f91d82d9947d97bfb

## Non-standard Abbreviations and Acronyms

SRCR: Swedish Registry of Cardiopulmonary Resuscitation
EMDC: Emergency Medical Dispatch Center
OHCA: Out-of-hosptial cardiac arrest
PSAP: Public safety answering point
EMS: Emergency Medical Services
CPR: Cardiopulmonary resuscitation
SOS alarm: 14 out of 18 regional EMDC
SvLc: Sjukvårdens larmcentral, 4 out of 18 regional EMDC.
AHA: American Heart Association
CrI: Credible interval

